# Impact of Targeted Therapy on Progression-Free Survival in Breast Cancer: A Decade of Evidence from Randomized and Clinical Trials

**DOI:** 10.1101/2025.10.31.25339265

**Authors:** Oladipo Gideon Adesina, Femi Nwagwuogbe

**Author notes:** **Corresponding Author Oladipo Gideon Adesina**, University of Benin, Faculty of Pharmacy, University of Benin, Benin City, Edo State, Nigeria. **Ethical Conformity Statement** This article is a systematic review of published studies and did not involve the direct participation of human subjects. As such, ethical approval and informed consent were not required.

## Abstract

**Background:** Targeted therapies have transformed breast cancer management by focusing on molecular drivers such as HER2 amplification, hormone receptor signaling, and CDK4/6 pathways. They aim to increase disease control while reducing systemic toxicity compared to conventional chemotherapy.

**Objective:** This review assessed the effectiveness and safety of targeted therapies in advanced and metastatic breast cancer, with progression-free survival (PFS) as the primary outcome and overall survival (OS), objective response rate (ORR), and safety as secondary endpoints.

**Methods:** A systematic search of PubMed/MEDLINE and the Cochrane Library was conducted between January 2015 to March 2025 for randomized controlled trials and large clinical studies comparing conventional and targeted pharmacotherapy in breast cancer. Eligible studies reporting progression-free survival were included. PRISMA guidelines were strictly followed. Data extraction and risk-of-bias assessments were performed independently by two reviewers.

**Results:** Fifteen trials with more than 10,000 patients were included. In HER2-positive disease, trastuzumab deruxtecan (T-DXd) significantly improved PFS and OS compared with trastuzumab emtansine, while tucatinib combinations provided strong intracranial control. CDK4/6 inhibitors (ribociclib, palbociclib, abemaciclib) consistently extended PFS in HR+/HER2-populations across pre- and postmenopausal groups. For triple-negative breast cancer, atezolizumab plus nab-paclitaxel improved outcomes in PD-L1-positive patients. Safety profiles were distinct, with interstitial lung disease from T-DXd, hematologic toxicity from CDK4/6 inhibitors, and immune-related events with checkpoint inhibitors.

**Conclusions:** Targeted therapies substantially improve PFS, with OS benefits in several trials, setting new standards across breast cancer subtypes. Balancing efficacy with toxicity management and improving global access remain essential to maximize clinical benefit.

## INTRODUCTION

Breast cancer is the most common cancer in women all over the world and a significant cause of cancer-related death. In 2020, there were estimated 2.3 million new cases and 685,000 deaths.^1^ Even with improvements in early detection and therapy, metastatic breast cancer is still a significant clinical issue. Cures are rare, and survival varies depending on the cancer subtype and access to healthcare.^2^ Systemic therapy in the past was based on cytotoxic chemotherapy, hormone therapy in the case of hormone receptor-positive disease, and HER2-directed monoclonal antibodies. While these treatments yielded benefits, they were all subject to compromise by resistance ^3^, reducing long-term effectiveness and leading to sequential therapy.^4^ This drawback encouraged the development of more targeted therapies created to increase effectiveness while minimizing systemic side effects.

Targeted therapies differ from conventional chemotherapy as they attack unique weaknesses in cancer cells rather than attacking all dividing cells.^5^ In breast cancer, they include inhibiting the amplification of HER2, manipulating estrogen receptor signaling, and blocking the CDK 4/6 pathway.^6^ Contrary to conventional chemotherapies, they provide longer-lasting responses and fewer systemic side effects.^7^ Whereas this shift has brought new challenges to cancer therapies, it has also imposed new demands. Such agents can potentially cause particular toxicities, for instance, cardiac, hepatic, or pulmonary toxicity, which must be watched closely and assessed over the long term for safety.^8^ Thus, the greatest challenge of modern breast cancer therapy is how to balance the benefits of targeted therapy with its risks.

Progression-free survival (PFS) now constitutes an essential measure of efficacy for such therapies. Though OS is the ultimate endpoint, it is influenced by crossover therapy, many other therapies, and long times to survival that conceal differences between regimens.^9^ PFS provides an earlier and more accurate impression of benefit from treatment since it indicates to what extent therapy delays progression.^10^ This is most marked for targeted therapy, which has been able to control disease prior to the emergence of differences in OS.^7^ In terms of safety, PFS also quantifies how long patients are on the drug and its toxicities, making it a fair endpoint.^5^

The pace of drug development in breast cancer has accelerated, resetting standards of care between subtypes. In HER2-positive cancer, antibody-drug conjugates and HER2 tyrosine kinase inhibitors are alternatives to chemotherapy and trastuzumab.^11^ In HR+/HER2-negative cancer, CDK4/6 inhibitors revolutionized endocrine therapy and improved disease control.^12^ This advancement points towards a shift in the field of oncology towards biologically targeted treatment.^13^ But they also create pragmatic concerns about sequencing, combination administration, and overlap toxicities.^14^ Resistance remains inevitable, cost restricts access, and extended safety monitoring is required.^4,8,14^ Thus, while targeted therapies have transformed outcomes, sustainability and equity issues remain.

By the integration of evidence from high-level clinical trials and observational studies, the review aims to present clinicians, researchers, and policymakers with an informed and balanced view of targeted therapy in breast cancer, with new treatment strategies enhancing effectiveness, safety, and accessibility.

## METHODS

The systematic review adhered to the Preferred Reporting Items for Systematic Reviews and Meta-Analyses (PRISMA) guideline, which provides a stepwise framework to transparent and reproducible reporting.^15^ The protocol was established in advance to ensure methodological consistency, including eligibility criteria, search strategy, study selection, data extraction, and bias assessment. Two separate reviewers performed all review stages to prevent bias and ensure reliability, with differences resolved by discussion till consensus.^17^

### Population

The review was focused on adult breast cancer patients, either by site or stage, with the particular focus on advanced and metastatic disease where progression-free survival (PFS) is especially relevant.

### Intervention

Interventions considered for use were targeted therapies such as HER2-targeted monoclonal antibodies, antibody–drug conjugates, HER2 tyrosine kinase inhibitors, CDK4/6 inhibitors, and immunotherapies that are different from conventional chemotherapy.

### Comparisons

Comparisons consisted of standard systemic therapies such as routine chemotherapy regimens, hormonal therapy alone without the use of targeted therapy, or other pre-targeted systemic therapy.

### Outcomes

The primary outcome of interest was PFS, as measured in months or hazard ratios. Secondary endpoints were overall survival (OS), objective response rates (ORR), and toxicity profiles because they matter in clinical interpretation.

### Study design

Randomized controlled trials (RCTs), cohort studies, and prospective clinical trials were taken into account. Case reports, case series, preclinical information, and incomplete conference abstracts were excluded to maintain reliability.

### Study period

Studies from January 2015 through March 2025 were included. This period encompasses the advent of CDK4/6 inhibitors, antibody–drug conjugates such as trastuzumab deruxtecan, and HER2 tyrosine kinase inhibitors such as tucatinib. The first half of this time frame covers conventional chemotherapy and the early years of trastuzumab treatment, and later years reflect the rapid expansion of biologically targeted therapies.^6^ Restricting the analysis to this timeframe maximizes clinical relevance to current-day oncology treatment and guidelines.

### Information sources and search strategy

Thorough searches were conducted in PubMed/MEDLINE and the Cochrane Library, the most trustworthy databases for oncology research.^16^ The search strategy was progressively optimized using free-text words and Medical Subject Headings (MeSH), combined with Boolean operators. Terms employed included disease descriptors (“breast cancer,” “breast carcinoma,” “breast neoplasm”), therapy classifications (“pharmacological therapy,” “chemotherapy,” “endocrine therapy,” “hormonal therapy”), targeted therapies (“HER2-targeted therapy,” “trastuzumab,” “pertuzumab,” “CDK4/6 inhibitors,” “immunotherapy,” “monoclonal antibodies”), and outcomes of interest (“progression-free survival,” “PFS”). Results were restricted to RCTs, clinical trials, and observational cohort studies. Reference lists of included articles were manually searched for other relevant studies.

### Study selection

Duplicates were removed using Zotero (version 1.0.54), which efficiently screens between databases.^18^ Independent screening of the abstracts and titles against inclusion criteria, and subsequent full-text evaluation of potentially eligible studies, was performed by two reviewers. Disagreements were resolved collaboratively, and consensus through discussion was attained, according to Cochrane standards.^17^

### Data extraction

A standard pre-defined form was used to extract study design, sample size, population, intervention and comparator description, outcome measures, follow-up duration, and reported PFS outcomes. Extraction was performed by two reviewers separately to minimize error. In the case of multiple published studies, the most extensive dataset was used; authors were contacted to clarify where necessary.

### Risk of bias assessment

Risk of bias was assessed on the basis of study design. For RCTs, the Cochrane Risk of Bias tool version 2 (RoB 2) was employed, which included judging randomization, deviations from the planned interventions, missing data on outcomes, measurement of the outcome, and reporting bias.^19^ For non-randomized studies, the Newcastle-Ottawa Scale (NOS) was utilized, which has wide validation for assessing cohort and case-control studies in oncology. NOS was selected over tools such as ROBINS-I because it is simple, with its confirmed usage in clinical studies, and suitability for the large number of observational designs included.^20^ The two reviewers independently evaluated every study, arriving at a consensus in case of disagreement.

### Data synthesis

Both the quantitative and qualitative procedures were employed to synthesize the data. Where study designs were sufficiently similar in terms of interventions, comparators, and outcomes, meta-analysis was conducted using a random-effects model. Hazard ratios for PFS were meta-analyzed where available, with heterogeneity explored using the I^2^ statistic. Where pooling was not feasible, evidence was synthesized narratively, by breast cancer subtype, intervention type, and comparator. Pre-defined sensitivity analyses to examine the impact of excluding studies at high risk of bias were investigated.

## RESULTS

812 studies were identified through PubMed/MEDLINE and the Cochrane Library. The removal of 176 duplicates in Zotero gave 636 unique records that were title and abstract screened. 94 full-text articles were reviewed and 15 trials were considered to fulfill the inclusion criteria. The main exclusion criteria were the lack of PFS data, non-comparative study, or study types outside the purview of this review. The selection of studies is illustrated in the PRISMA flow diagram (Figure 1). Characteristics of the included studies are shown in Table S1 (supplementary table). Of the 15 included trials, published between 2015 and 2024, they were phase II and phase III RCTs and large multicenter phase IIIb trials. Altogether, they enrolled more than 10,000 patients with advanced or metastatic breast cancer. Sample sizes ranged from 184 patients in discovery trials to over 2,888 in multinational large studies. Trials were conducted in North America, Europe, and Asia, thereby improving generalizability across populations. Follow-up periods ranged from 18 months to over five years, enabling both short- and long-term measurement of outcomes.

**Figure 1.**
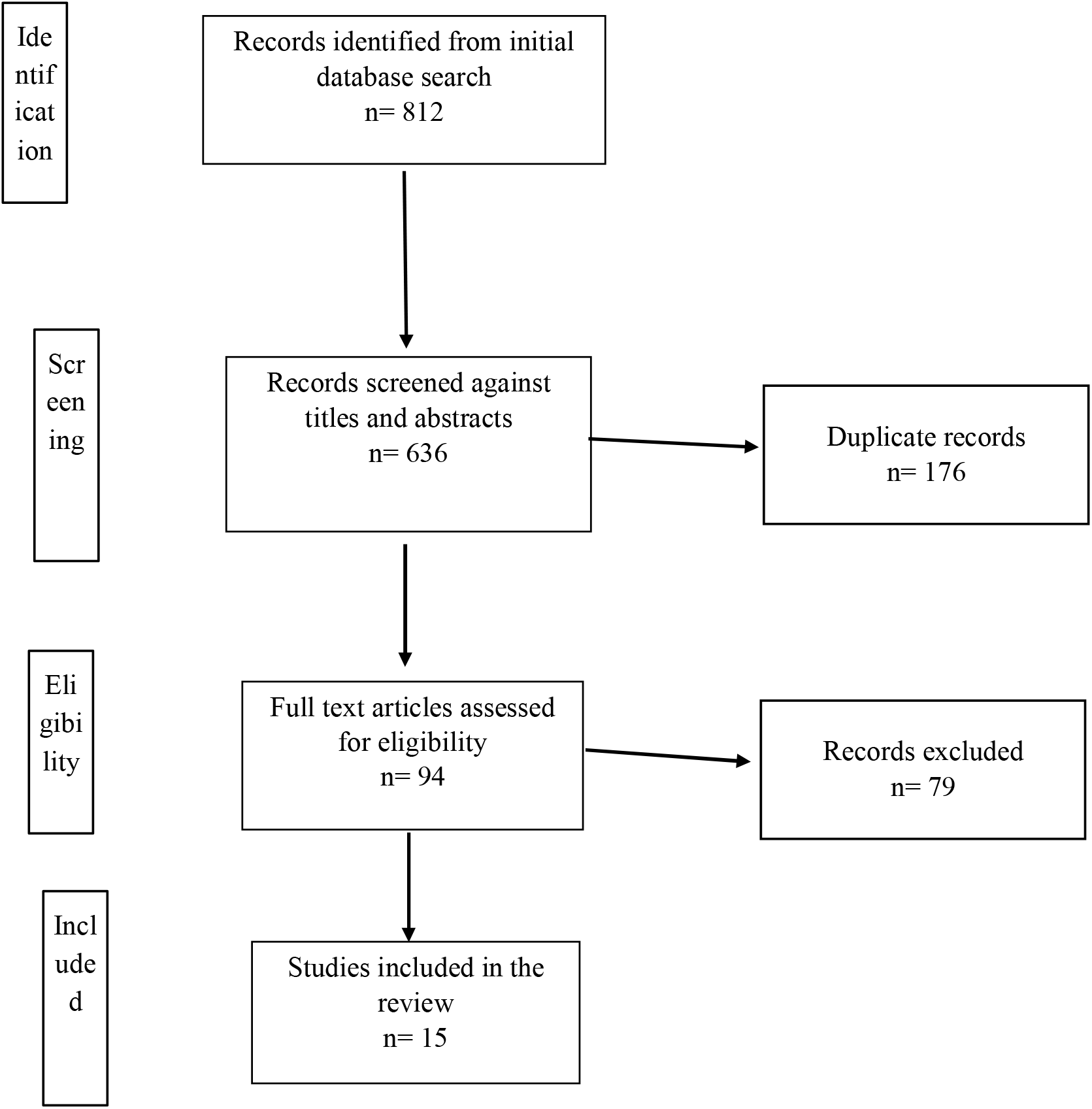
PRISMA flowchart showing selection of studies.

HER2-positive disease was dominant in the dataset. Trials compared consecutive generations of HER2-directed treatments, including among them antibody-drug conjugates such as trastuzumab deruxtecan in trastuzumab and chemotherapy-treated patients. Tyrosine kinase inhibitors such as tucatinib were explored with capecitabine and trastuzumab, including subgroups in central nervous system disease. Pertuzumab and trastuzumab with taxane skeletons were other first-line regimens investigated. Trastuzumab emtansine, chemotherapy, or placebo with standard HER2-directed agents were comparators. Together, these trials illustrate an evolving trend where more recent HER2-directed medications follow upon failure of earlier regimens.

Hormone receptor-positive, HER2-negative disease was treated in several large RCTs of cyclin-dependent kinase (CDK) 4/6 inhibitors (ribociclib, palbociclib, abemaciclib). They were compared with aromatase inhibitors or fulvestrant, versus endocrine therapy alone. Most were initial treatment studies in postmenopausal women with advanced disease. Their long sample sizes and long follow-up generated high-quality evidence of substantial PFS benefit over standard endocrine therapy.

Triple-negative breast cancer and immunotherapy were underrepresented, but there are reported studies that are recognized for having supplied evidence for expanding checkpoint inhibitor use in PD-L1–positive disease. Although fewer in number, they expand the therapeutic landscape and are a testament to ongoing investigation of biomarker-targeting treatment.

Study quality and design were heterogeneous. Most were randomized and controlled, but a few used open-label designs. Open-label designs carry risks of performance and detection bias, but PFS was always assessed on standardized criteria and often confirmed by blinded central assessment. Single-arm trials were used as phase II exploratory studies in some cases. In all trials, PFS was typically a primary or prespecified secondary endpoint and therefore could be compared.

Risk of bias assessment illustrated overall high methodological quality. Randomized trials had low risk of randomization and allocation concealment. Open-label trials had moderate performance/detection bias, which was mitigated by blinded outcome assessment. Attrition was minimal with fewer than 10% missing outcome data in most trials. Crossover between treatment groups made PFS and OS interpretation problematic in some cases. Observational and single-arm trials had lower scores according to design limitations but provided supportive background. Safety and adverse effects were commonly reported. HER2-specific antibodies–drug conjugates had pulmonary toxicities such as interstitial lung disease (10–15% in bigger trials). CDK4/6 inhibitors were more commonly linked with hematologic toxicities like neutropenia and isolated hepatotoxicity. Tucatinib was linked with gastrointestinal side effects and transaminase elevation. Comparators with chemotherapy experienced more nonspecific adverse events like alopecia, fatigue, and gastrointestinal side effects. Grade 3 or higher toxicities were commonly reported, which facilitated fair risk–benefit analysis.

## DISCUSSION

Targeted therapy has transformed the treatment of breast cancer subtypes, consistently improving progression-free survival (PFS) compared with standard of care and, in many trials, extending overall survival (OS).^21-25^ The most powerful effects were in HER2-positive illness, wherein antibody–drug conjugates such as trastuzumab deruxtecan (T-DXd) decisively beat trastuzumab emtansine (T-DM1) in DESTINY-Breast03 with a progression or death hazard ratio of 0.28 and an objective response rate of near 80%,^21^ whereas interim findings from DESTINY-Breast01 provided a median PFS of 19.4 months with aggressive pretreatment.^8^ Small-molecule inhibitors such as tucatinib have also redirected treatment in brain-metastatic cohorts, with HER2CLIMB extending systemic PFS from 5.6 to 7.8 months and approximately doubling intracranial PFS.^23,24^ At the same time, cyclin-dependent kinase 4/6 (CDK4/6) inhibitors have become standard for hormone receptor–positive (HR+), HER2-negative breast cancer, prolonging PFS in both the postmenopausal and premenopausal populations with the use of endocrine therapy.^6,12,13,26,27^ For TNBC, the addition of checkpoint blockade with atezolizumab and nab-paclitaxel improved PFS from 5.0 to 7.5 months in PD-L1-positive disease (HR 0.62), establishing a biomarker-guided approach in a setting that had previously relied solely on chemotherapy.^28^ Beyond randomized controlled trials, comparative real-world evidence corroborates that these benefits persist in larger clinical populations,^29^ and single-arm trials and patient-reported outcomes data, though less definitive, indicate durability and quality-of-life benefits consistent with randomized findings.^8,25,30^

In HER2-positive, T-DXd delivers the most conclusive PFS benefit, soundly defeating T-DM1 in DESTINY-Breast03 with a 0.28 risk ratio of progression or death and objective response rate of about 80%.^21^ Follow-up analysis established an actual OS benefit, setting it as first-line gold standard following trastuzumab and pertuzumab.^22^ Prior single-arm experience in DESTINY-Breast01 demonstrated similar durabilities with median PFS of 19.4 months and median OS of 29.1 months despite intensive pretreatment.^8^ More importantly, PRO-based findings indicate that quality of life was maintained even with prolonged exposure, thereby endorsing its clinical value.^30^ However, pneumonitis and interstitial lung disease, presented in 10–16% of patients, are significant threats and require close watch.^8,21,22^ Combined, these results establish T-DXd as the strongest systemic therapy for HER2-positive metastatic disease and reinforce the necessity of systematic toxicity surveillance.

Tucatinib-based treatment have addressed an unmet need for patients with CNS involvement. HER2CLIMB improved systemic PFS from 5.6 to 7.8 months and approximately doubled intracranial PFS in brain metastasis patients, with OS benefit extending to this subgroup.^23^ The consistency of these results across endpoints was reinforced by a companion analysis.^24^ Compared to antibody-drug conjugates, tucatinib has a distinct side effect profile, including transaminase elevations and diarrhea, which are manageable and allow for individualized treatment approaches based on disease distribution.^23,24^ By contrast, first-line real-world evidence for pertuzumab, trastuzumab, and a taxane have reported PFS of over 20 months, although randomization limitation limits high-end comparative analysis.^31^ These results suggest treatment strategy with dual-antibody therapy remaining the cornerstone first-line, utilizing T-DXd in systemic disease and tucatinib combinations in intracranial disease, with choices dependent on site of disease and tolerance.^21-25^

HER2-low breast cancer, defined by non-amplification with low receptor expression, expands the window for targeted therapy. In DESTINY-Breast04, T-DXd halved the risk of progression versus physician’s-choice chemotherapy with a median PFS of 10.1 months versus 5.4 months (HR 0.50; 95% CI 0.40–0.63) and a significant overall survival advantage of 23.4 months versus 16.8 months.^31^ This groundbreaking finding reclassifies HER2 as a spectrum rather than as a binary marker and emphasizes the importance of uniform pathology assessment to identify suitable patients. The same risk of interstitial lung disease associated with HER2-positive disease is present here, with the need to recognize it early on and treat it urgently to preserve benefit.^31^ For HR+/HER2-negative disease, CDK4/6 inhibitors have transformed expectations by significantly prolonging PFS when added to endocrine therapy. PALOMA-2 proved that addition of palbociclib to letrozole improved PFS to 24.8 months vs 14.5 months on letrozole alone, but OS remained nonsignificant, which can be explained by following successful therapy^13.^ MONARCH 2 and MONARCH 3 extended this benefit to abemaciclib, with OS benefits on extended follow-up appearing, particularly in endocrine-resistant patients.^27^ Ribociclib has produced the strongest evidence to this point, with MONALEESA-2 showing strong PFS and follow-up OS benefit of about 12 months, and MONALEESA-7 showing comparable gains in premenopausal and perimenopausal women, a hitherto under-represented population.^6,12,26^ Observational real-world data from P-REALITY X validated the greater PFS and OS with palbociclib and aromatase inhibitor compared to aromatase inhibitor monotherapy in clinical practice, in support of external validity.^29^ In these trials, diarrhea and hematologic toxicities were common but dose-adjustable, allowing for treatment to be prolonged and PFS advantage to be gained.^6,12,13,27^

In TNBC, in the IMpassion130 trial, atezolizumab plus nab-paclitaxel greatly favored PD-L1– positive disease, with median PFS 7.5 months compared with 5.0 months with nab-paclitaxel alone (HR 0.62; 95% CI 0.49–0.78), cementing the role for immune checkpoint blockade in this high-risk subset.^28^ Overall survival (OS) outcomes were more heterogeneous, with the result that PFS is no longer necessarily a reliable surrogate with patients having access to multiple subsequent therapies. Immunotoxicities such as pneumonitis, colitis, and endocrinopathies did occur, but these are usually manageable with early steroid treatment and thus active monitoring is important. The trial did however firmly establish immunotherapy as an integral part of front-line treatment in biomarker-positive TNBC.

Supportive background is derived from nonrandomized trials and PRO analysis. PERUSE trial reported a PFS median of 20.7 months with pertuzumab, trastuzumab, and taxane, in accordance with randomized data, also confirming paclitaxel as an alternative backbone.^25^ DESTINY-Breast01 exhibited durable response with T-DXd in heavily pretreated patients, foreshadowing the superiority results later confirmed over T-DM1.^8,21^ PROs from DESTINY-Breast02 have shown quality of life is maintained on T-DXd, reflective of its tolerability with disease control prolonging survival.^30^ While not a replacement for randomized trials, their agreement with RCT data enhances confidence in the representativeness of targeted therapy.^8,25,30^

Safety cannot be ignored alongside efficacy. T-DXd carries a high risk of interstitial lung disease; tucatinib is susceptible to liver and gastrointestinal toxicities; CDK4/6 inhibitors cause predictable neutropenia and diarrhea; and immune checkpoint inhibitors can lead to serious immune-mediated events.^13,21-24,27-28^ In spite of these toxicities, they are most often managed by observation, dose adjustment, and prompt intervention, allowing for extended therapy and maximization of PFS benefit. This trade-off between effectiveness, represented as PFS gains, versus tolerability, is the value-proposition of targeted therapy.

Uncertainties remain regarding sequencing and access. For HER2-positive illness, T-DXd and tucatinib sequencing is reasonable but head-to-head sequencing trials are lacking.^21-25^ CDK4/6 inhibitors are standard initial treatment in HR+ disease, but optimal practice on progression, class switch, combination with other endocrine partners or inhibition of new pathways differs by resistance biology and tolerance in patients.^6,12,13,26,27^ Access gaps, reimbursement limits, and infrastructure hurdles make cost the key barrier to equitable benefit.^29,32^ These structural impediments are as important as trial efficacy in defining population-level outcomes.

## CONCLUSION

The overall combined set of information for fifteen trials indicates that targeted therapies are linked with improved progression-free survival compared with standard treatment in breast cancer, with several regimens also accruing overall survival advantage. The biggest benefit is in HER2-positive disease, where trastuzumab deruxtecan and tucatinib regimen have become standard for systemic and brain treatment, and HER2-low tumors now have options with antibody-drug conjugates. For HR-positive, HER2-negative breast cancer, CDK4/6 inhibitors have set a new standard with significant improvement in disease control in postmenopausal and premenopausal groups. In triple-negative disease, immunotherapy combination with checkpoint inhibitors has demonstrated a biomarker-directed strategy that slows decline in PD-L1–positive disease. In each of these subtypes, the magnitude of the PFS gain is linear and constant, and targeted therapy thus forms the basis of modern breast cancer therapy.

These gains are still unequal. Some trials, such as PALOMA-2 and IMpassion130, had imperfect or variable PFS correlation with overall survival, highlighting that PFS is not a surrogate endpoint for everyone. Toxicities are similarly distinct and clinically relevant, ranging from interstitial lung disease with trastuzumab deruxtecan to hematologic suppression with CDK4/6 inhibitors and immune-related toxicities with checkpoint blockade. Those safety issues require close monitoring and prompt intervention. Furthermore, trial sites are usually locations in high-income nations, and concerns for global equity arise due to expensive drugs and infrastructural constraints reducing access in low- and middle-income contexts. Therefore, as new targeted treatments have revolutionized breast cancer therapy, its full potential will only be reached with simultaneous consideration of efficacy, safety, and access.

## Data Availability

All data produced in the present work are contained in the manuscript.

## RECOMMENDATIONS

Subsequent sequencing trials would be helpful to establish the most optimal sequence of targeted therapy, for example, in HER2-positive disease where antibody-drug conjugates and small-molecule inhibitors are both effective but not compared head-to-head. Optimization of biomarkers is necessary. HER2-low status should enter routine diagnostic practice, and assessment of resistance in HR-positive disease determines if patients might benefit from switching of CDK4/6 inhibitors, or moving to new targeted pathways. Checkpoint blockade in TNBC requires additional predictive indicators, beyond PD-L1 status, to identify patients who will benefit long-term.

Clinical care should integrate patient-reported outcomes into routine surveillance to ensure that extended progression-free survival aligns with sustained quality of life. Toxicity management standards should be institutionalized, early identification of interstitial lung disease, preemptive treatment of neutropenia, and immune-related adverse event guidelines should be implemented within care. Real-world evidence should continue to validate trial effectiveness in larger populations and inform adaptation where resource limitations limit access.

Policy and health systems response are also important. Antibody-drug conjugates and chronic oral targeted agents are expensive, and enhanced access to biosimilars, rate rebalancing, and access to target agents on essential medicines lists to improve access are required. Investment in diagnostic infrastructure, including accessible and reproducible immunohistochemistry for HER2-low and PD-L1 testing, is also required to promote equitable access. Short of such interventions, the gains in progression-free survival demonstrated in clinical trials will remain limited to a minority of patients.

## ABBREVIATIONS

ADC: Antibody–drug conjugate
CDK4/6: Cyclin-dependent kinase 4/6
HER2: Human epidermal growth factor receptor 2
HR+: Hormone receptor–positive
mBC: Metastatic breast cancer
NGS: Next-generation sequencing
ORR: Objective response rate
OS: Overall survival
PFS: Progression-free survival
PRISMA: Preferred Reporting Items for Systematic Reviews and Meta-Analyses
PRO: Patient-reported outcomes
RCT: Randomized controlled trial
TNBC: Triple-negative breast cancer
T-DM1: Trastuzumab emtansine
T-DXd: Trastuzumab deruxtecan

## Notes

**Conflict of Interest Declaration** The authors declare no conflicts of interest related to this work.

### Competing Interest Statement

The authors have declared no competing interest.

### Funding Statement

This study did not receive any funding.

