## Supplementary material for "Impact of Targeted Therapy on Progression-Free Survival in Breast Cancer: A Decade of Evidence from Randomized and Clinical Trials": Table S1 (supplementary table)

### TABLE S1: CHARACTERISTICS OF THE STUDY

| Name | Design | Population | N | Treatment | Objectives | Results | Follow-up |
| --- | --- | --- | --- | --- | --- | --- | --- |
| Cortés et al., DESTINY-Breast03 (2022) | Phase 3, open-label, randomized | HER2+ metastatic breast cancer, previously treated with trastuzumab + taxane | 524 | T-DXd vs T-DM1 | Compare progression-free survival (PFS) | 12-mo PFS: 75.8% vs 34.1%; ORR 79.7% vs 34.2%; OS trend favored T-DXd | Median ~28 months at primary; updated ~41 months |
| Fehm et al., DESTINY-Breast02 (2024) | Phase 3, randomized, open-label | HER2+ metastatic breast cancer, progressed on T-DM1 | 608 | Trastuzumab deruxtecan vs physician’s choice (capecitabine+trastuzumab or capecitabine+lapatinib) | Compare PFS (primary), OS (secondary); patient-reported outcomes | PFS 17.8 vs 6.9 mo (HR 0.36); OS 39.2 vs 26.5 mo (HR 0.66). PROs favored T-DXd | Median ~21.5 months |
| Finn et al., PALOMA-2 (2016) | Phase 3, randomized, double-blind | Postmenopausal HR+/HER2– advanced breast cancer, untreated | 666 | Palbociclib + letrozole vs placebo + letrozole | Confirm benefit of adding palbociclib | PFS 24.8 vs 14.5 mo (HR 0.58); higher myelotoxicity with palbociclib | Median 23 months |
| Harbeck et al., MONALEESA-7 QoL (2020) | Phase 3, randomized | Premenopausal HR+/HER2– advanced breast cancer | 672 (335 ribociclib, 337 placebo) | Ribociclib + endocrine therapy vs placebo + endocrine therapy | Evaluate quality of life alongside PFS/OS | Ribociclib delayed QoL deterioration (HR 0.67); also improved PFS 23.8 vs 13.0 mo, OS HR 0.71 | Median 42 months |
| Hortobagyi et al., MONALEESA-2 (2016) | Phase 3, randomized, double-blind | Postmenopausal HR+/HER2– advanced breast cancer, untreated | 668 | Ribociclib + letrozole vs placebo + letrozole | Assess PFS and OS | PFS HR 0.56; 18-mo PFS 63% vs 42%; ORR 52.7% vs 37.1% | Median 15.3 months (long-term OS updated later) |
| Hortobagyi et al., MONALEESA-2 OS final (2022) | Phase 3, randomized, double-blind | Postmenopausal women with HR+/HER2– advanced breast cancer | 668 | Ribociclib + letrozole vs placebo + letrozole | Overall survival (secondary) | OS: 63.9 vs 51.4 mo (HR 0.76) | Median follow-up 6.6 years |
| Hurvitz et al., DESTINY-Breast03 updated (2023) | Phase 3, randomized, open-label | HER2+ metastatic breast cancer, previously treated with trastuzumab + taxane | 524 | Trastuzumab deruxtecan vs trastuzumab emtansine | Compare PFS (primary), OS (secondary) | PFS 28.8 vs 6.8 mo (HR 0.33); OS HR 0.64 | Median ~28 months |
| Lin et al., HER2CLIMB exploratory brain mets (2020) | Exploratory analysis of Phase 2 HER2CLIMB | HER2+ metastatic breast cancer with active/stable brain metastases | 291 | Tucatinib regimen vs placebo regimen | CNS-PFS, OS | CNS-PFS HR 0.32; median OS 18.1 vs 12.0 mo | Median follow-up ~29.6 months |
| Miles et al., PERUSE (2021) | Phase IIIb, single-arm, multicentre | HER2+ locally recurrent/metastatic breast cancer, untreated in advanced setting (except endocrine) | 1,436 | Pertuzumab + trastuzumab + taxane (docetaxel, paclitaxel, or nab-paclitaxel) | Assess safety; explore PFS/OS | Median PFS 20.7 mo; OS 65.3 mo. Outcomes similar across taxanes | Median 5.7 years |
| Modi et al., DESTINY-Breast04 (2022) | Phase 3, randomized | HER2-low metastatic breast cancer, 1–2 prior chemo lines | 557 | T-DXd vs physician’s choice chemo | PFS in HR+ group (primary) | PFS: 10.1 vs 5.4 mo (HR 0.51); OS: 23.9 vs 17.5 mo | Median follow-up ~18.4 months |
| Murthy et al., HER2CLIMB (2020) | Phase 2, randomized, double-blind | HER2+ metastatic breast cancer, prior trastuzumab, pertuzumab, and T-DM1; brain mets allowed | 612 | Tucatinib + trastuzumab + capecitabine vs placebo combo | PFS (primary), OS (secondary) | PFS HR 0.54; OS HR 0.66; brain mets PFS HR 0.48 | Median ~29.6 months |
| Rugo et al., Palbociclib RWE (2022) | Real-world comparative cohort | HR+/HER2− metastatic breast cancer starting first-line therapy | 2,888 | Palbociclib + aromatase inhibitor vs aromatase inhibitor | Compare overall survival and time to progression in routine care | OS 49.1 vs 43.2 mo, HR 0.76; rwPFS 19.3 vs 13.9 mo, HR 0.70 | Cutoff Sep 30, 2020; ≥6-month potential follow-up |
| Saura et al., DESTINY-Breast01 (2024 update) | Phase 2, single-arm | Heavily pretreated HER2+ metastatic breast cancer, prior T-DM1 | 184 | T-DXd | Objective response rate | ORR 62%; median OS 29.1 mo; PFS 19.4 mo | Median 26.5 months |
| Schmid et al., IMpassion130 (2018) | Phase 3, randomized, double-blind | Untreated metastatic triple-negative breast cancer | 902 (451/arm) | Atezolizumab + nab-paclitaxel vs placebo + nab-paclitaxel | Assess PFS and OS | PFS 7.2 vs 5.5 mo overall; 7.5 vs 5.0 mo in PD-L1+. OS 25.0 vs 15.5 mo in PD-L1+ | Median 12.9 months |
| Sledge et al., MONARCH-2 (2017) | Phase 3, randomized, double-blind | HR+/HER2– advanced breast cancer, progressed on prior endocrine therapy | 669 | Abemaciclib + fulvestrant vs placebo + fulvestrant | Assess PFS (primary), OS (secondary) | PFS 16.4 vs 9.3 mo (HR 0.55); ORR 48% vs 21% | Median ~27 months |
